# Life-course exposure to air pollution and biological ageing in the Lothian Birth Cohort 1936

**DOI:** 10.1101/2022.04.17.22273946

**Authors:** Gergő Baranyi, Ian J. Deary, Daniel L. McCartney, Sarah E. Harris, Niamh Shortt, Stefan Reis, Tom C. Russ, Catharine Ward Thompson, Massimo Vieno, Simon R. Cox, Jamie Pearce

## Abstract

**Background:** Exposure to ambient air pollution is associated with a range of diseases. Biomarkers derived from DNA methylation (DNAm) indicate a potential pathway to human health differences, connecting disease pathogenesis and biological ageing. However, little is known about sensitive periods during the life course where air pollution might have a stronger impact on DNAm, or whether effects accumulate over time.

**Objectives:** We examined associations between air pollution exposure across the life course and DNAm-based biomarkers of ageing.

**Methods:** Data were derived from the Lothian Birth Cohort 1936. Participants’ residential history was linked to annual levels of PM_2.5_, SO_2_, NO_2_, and O_3_ around 1935, 1950, 1970, 1980, 1990, and 2001; pollutant concentrations were estimated using the EMEP4UK atmospheric chemistry transport model. Blood samples were obtained between ages of 70 and 80 years, and Horvath DNAmAge, Hannum DNAmAge, DNAmPhenoAge, DNAmGrimAge, and DNAm telomere length (DNAmTL) were computed. We applied the structured life-course modelling approach: least angle regression identified best-fit life-course models for a composite measure of air pollution (air quality index [AQI]), and mixed-effects regression estimated selected models for AQI and single pollutants.

**Results:** We included 525 individuals with 1782 observations. In the total sample, increased air pollution around 1970 was associated with higher epigenetic age (AQI: b=0.622 year, 95%CI: 0.151, 1.094) measured with Horvath DNAmAge in late adulthood. We found shorter DNAmTL among males with higher air pollution around 1980 (AQI: b=-0.035 kilobase, 95%CI: -0.057, -0.014) and among females with higher exposure around 1935 (AQI: b=-0.036 kilobase, 95%CI: -0.059, -0.013). Findings passed false discovery rate correction for DNAmTL, and were more consistent for the pollutants PM_2.5_, SO_2_ and NO_2_.

**Discussion:** We tested the life-course relationship between air pollution and DNAm-based biomarkers. Air pollution *in utero* and in young-to-mid adulthood is linked to accelerated epigenetic ageing and telomere-associated ageing in later life.

## INTRODUCTION

Ambient air pollution is one of the greatest environmental threats to human health, with serious consequences for morbidity and mortality,^1^ and is responsible for an estimated $3.5 trillion welfare and $144 billion labour income losses annually worldwide.^2^ Based on the Global Burden of Diseases Study 2015, 4.2 million annual deaths and 103 million disability-adjusted life years can be attributed to exposure to fine particulate matter with an aerodynamic diameter of ≤2.5 µm (PM_2.5_).^3^ Gaseous pollutants, such as nitrogen dioxide (NO_2_), sulphur dioxide (SO_2_) and ozone (O_3_) have been also associated with hazardous health effects and increased mortality.^1^ The burden is largest in low-and middle-income countries; however, levels also exceed WHO air quality standards^1^ in high-income countries, with even low-level exposures being associated with increased mortality.^4^ Research has established the impact of long-term exposure to air pollution on cardiovascular and respiratory diseases, and cancer,^3, 5^ and there is growing evidence for its effect on the risk of neurodegenerative^6^ and mental disorders.^7^

Air pollutants arriving in the lung can trigger multiple cellular mechanisms and translocate into the circulatory system; proposed biological pathways to adverse health outcomes include cytotoxicity, systematic inflammation and oxidative stress.^8^ Prolonged air pollution exposure can lead to leukocyte telomere length attrition (i.e. attrition of nucleoprotein complexes located at the ends of the chromosome),^9^ a marker of cellular senescence.^10^ There is also growing evidence highlighting the role of epigenetic alterations in response to environmental hazards, another hallmark of ageing.^10–12^ One key epigenetic mechanism that can contribute to the regulation of gene expression is DNA methylation (DNAm). DNAm describes the process of adding a methyl group to the fifth carbon of cytosine nucleotides in the genome, usually found at cytosine-phosphate-guanine (CpG) dinucleotides.^13, 14^ DNAm is susceptible to change: adverse exposures and stressors can modify DNAm patterns, and thus gene function and transcription, without altering the gene sequence.^13, 15^

Although appropriate epigenetic regulation is key for cell functioning, ageing cells undergo substantial changes in genome-wide DNA methylation levels, with specific genomic regions becoming hyper- or hypo-methylated.^16, 17^ Changes in methylation levels can be also observed in the development of various (age-related) diseases, including cancer.^18^ Given that accumulating alterations in DNA methylation patterns are associated with chronological age (i.e. calendar time since birth) across the life course,^19^ epigenetic age estimators (or epigenetic clocks) have been developed as markers of biological ageing representing the workings of epigenetic maintenance systems.^16, 17, 20^ Epigenetic clocks are derived from human tissues and calculated from the methylation level of sets of CpGs sites: whereas earlier clocks selected CpGs sites as best predicting chronological age, a surrogate of biological ageing, second generation clocks focus more on providing a prediction of healthy lifespan using other surrogate measures.^17^ Due to the strong correlation with chronological age, epigenetic age acceleration can be computed as the difference between chronological age and the epigenetic clock, with positive values indicating faster tissue ageing than expected.^17^ Accelerated ageing is a predictor of all-cause mortality^21^ and worse health outcomes,^17^ and it is associated with lower physical and cognitive fitness.^22^

Recent evidence indicates accelerated biological ageing, including telomere shortening and epigenetic alterations, among individuals exposed to long-term ambient air pollution (measured usually one year before outcome assessment)^9, 23, 24^; however, evidence is lacking on the relationship covering longer periods across the life course. It remains largely unknown whether there are sensitive or critical periods where exposure to air pollution has a prominent and/or long-lasting effect on biological ageing or – alternatively – whether the impact of air pollution gradually accumulates over time.^25^ These are profound questions as individuals’ ranking in leukocyte telomere length is relatively stable from birth to adulthood.^26^ Therefore, it is plausible that associations identified in later life^23, 27, 28^ are the consequence of exposures earlier in the life course combined with limited geographical mobility or moving to areas with similar air pollution levels.^29^ Prolonged air pollution exposure can often have a stronger impact on DNAm patterns than short-term exposure (a few days),^25^ which raises the question as to whether air pollution effects may accumulate over the life course.

The current study addresses this research gap by applying the life-course approach^30^ and exploring associations between air pollution from *in utero* onwards and DNAm-based biomarkers in late adulthood (i.e. epigenetic clocks, DNAm proxy of telomere length). Using a cohort of older, 1936-born Scottish adults with life-course residential addresses linked to historical air pollution concentrations and DNAm data in their 70s, we investigated whether local levels of pollution at different points across the life course (or their accumulation) are associated with biological ageing. Analyses focussed on four air pollutants (PM_2.5_, SO_2_, NO_2_, O_3_) with previously established impacts on health and wellbeing.^1^

## METHODS

### Study participants

We used data from the Lothian Birth Cohort 1936 (LBC1936).^31^ Participants were born in 1936 and took part in the Scottish Mental Survey 1947, a nationwide school-based cognitive test at the age of 11.^31^ Between 2004 and 2007, 1091 surviving men and women of the Scottish Mental Survey 1947 living in the City of Edinburgh and in the Lothian area of Scotland were retraced and recruited for participating in the LBC1936. The average age was 70 in the first wave. Follow-up waves took place at age ∼73 (2007-2010; n=866), ∼76 (2011-2013; n=697), and ∼79 (2014-2017; n=550).^31^ In 2014, LBC1936 participants were asked to complete a lifegrid questionnaire^32^ supported by ‘flashbulb’ memory prompts (e.g. 9/11 attacks in New York) aiming to capture residential histories from birth to the date of completing the lifegrid.^31^ Out of 704 individuals remaining in the study at that point, 593 participants provided 7423 addresses, which were georeferenced using automatic geocoders and historical building databases.^29^

### Historical air pollution exposure

Annual concentrations of PM_2.5_, SO_2_, NO_2_, and O_3_ were estimated using the EMEP4UK^33–35^ atmospheric chemistry transport model for the model years of 1935, 1950, 1970, 1980, 1990 and 2001 (details on data generation and the feasibility of using historical air pollution estimates in epidemiological research has been published previously)^6^. The model has a horizontal resolution of 0.5° × 0.5° used to provide the boundary condition for a nested UK domain with a horizontal resolution of 0.055° × 0.055° (∼5x6 km^2^). The EMEP4UK model have been extensively evaluated for the UK and globally.^36, 37^

Exposure to pollutants was derived based on latitudes and longitudes of geocoded residential addresses. We extracted annual concentrations of pollutants for each UK-based addresses using intervals around modelling years (i.e. 1935 [<1943], 1950 [1943-1959], 1970 [1960-1975], 1980 [1976-1985], 1990 [1986-1995], and 2001 [1996-2006]). Since participants could reside at multiple locations within a given time band, we calculated the mean level of exposure, resulting in no more than six estimates per pollutant for each participant.

Within each measurement period, exposure to pollutants was highly correlated: we found strong positive associations between PM_2.5_, SO_2_ and NO_2_ (Pearson correlation coefficients ranged between 0.58 and 1.00), and they were negatively correlated with O_3_ (ranged between -0.63 and -0.97) (Figure S1). Correlation coefficients were particularly high for the modelling years of 1935 and 1950. This is not surprising, given that during earlier years by far the largest emission sources were from coal/fossil fuel combustion, leading to high spatial and temporal correlation between PM_2.5_, SO_2_ and NO_2_; and the significant uncertainty of emissions for these modelling years. Weakening correlation over time might be explained by the introduction of clean air measures that particularly affected large, stationary combustion sources, and by road transport sources making a larger contribution.^38^

To reflect this high degree of shared variance between highly correlated air pollution exposures, we constructed a composite air quality index (AQI), an additive cumulative measure of multi-pollutants exposure.^39^ We first scaled and centred PM_2.5_, SO_2_, NO_2_ and O_3_ values, and then summed and scaled them for each measurement periods (i.e. AQI in 1935, 1950, 1970, 1980, 1990, and 2001); individual exposure to AQI was expressed in standard deviation (SD) changes.

### DNAm-based biomarkers

DNA methylation was derived from blood samples collected from LBC1936 participants in waves 1-4 (mean ages ∼70, 73, 76, 79 years). Methylation was measured at 485512 CpG sites using the Illumina HumanMethylation450BeadChips array; details are published elsewhere.^40^ Extensive quality control was carried out by removing (a) probes with low detection rate; (b) low-quality samples (e.g. inadequate hybridization); (c) samples with a low call rate (i.e. below 450000 probes); and (d) samples where sex based on XY probes did not match reported sex.^22^ After quality control, there remained 895 samples in wave 1, 792 in wave 2, 611 in wave 3 and 499 in wave 4.

Five biomarkers were computed from the available DNA methylation data. First generation clocks derived from chronological age included (1) Horvath’s multi-tissue epigenetic clock based on 353 CpGs,^19^ and (2) Hannum’s epigenetic clock based on 71 CpGs.^20^ We included also two second generation clocks: (3) for DNAm PhenoAge, CpGs were identified based on a composite measure of phenotypic age,^41^ and (4) DNAm GrimAge, a predictor of mortality, which incorporated DNAm surrogates for 7 plasma proteins and smoking pack-years.^42^ Finally, (5) a DNA methylation-based proxy of telomere length (DNAmTL) was derived based on 140 CpGs selected by regressing leukocyte telomere length on methylation data.^43^

### Covariates

Covariates are presented in a directed acyclic graph (DAG) taking into consideration the years of air pollution exposure and the timing of covariates during the life course (Figure S2). We considered age at the time of outcome assessment, sex (male, female) and parental occupational social class (OSC) (professional-managerial [I/II] versus skilled, partly skilled and unskilled [III/IV/V])^44^ as common confounders for all life-course hypotheses. Childhood smoking (initiating before the age of 16) was considered as a confounder from adolescence, years spent in full-time education from young adulthood onwards, whereas adult OSC (I/II versus III/IV/V),^44^ smoking status at age 70 (yes, no), and BMI at age 70 were considered as confounders during the second part of life.

### Statistical analysis

To explore the associations between exposure to air pollution across the life course and DNAm-based biomarkers in late adulthood, we applied the two-stage structured life-course modelling approach originally developed by Mishra et al^45^ and modified by Smith et al^46^ for continuous exposures. As different life-course models might be appropriate for biological ageing among males and females,^15, 28^ sex-stratified models are also presented. A flowchart outlining analyses can be found in Figure S3.

In the first stage, the life-course model(s) most strongly supported by the observed data were selected from multiple simultaneously competing ones by applying the least angle regression (LARS).^46^ LARS is a variable selection algorithm which implements the least absolute shrinkage and selection operator (lasso), and indicates the best lasso fit for each number of selected variables.^47, 48^ This approach always identifies the variable with the strongest association to the outcome in the observed data, and then it selects further variables based on their strength of association, applying an absolute value penalty; the process continues until all variables have been selected.^48^ In order to identify the most appropriate variable(s) supported by LARS, we used the covariance test for the lasso indicating whether additional variables significantly (p<0.05) improve explained outcome variance.^49^ Five biomarkers were tested in our study; to minimise type 1 errors arising from multiple comparisons, we provided false discovery rate (FDR) adjusted p-values for the covariance test (*p_FDR_*) across outcomes. Finally, as LARS cannot accommodate multilevel data structure with individuals having repeated outcome measurements, we conducted model selection for only one wave of DNAm-based biomarkers by choosing the wave with the largest sample size.

We investigated seven life-course models, which were inputted as variables into LARS. Sensitive periods captured exposure to air pollution based on the modelling years of 1935 (∼*in utero*), 1950 (∼14 years), 1970 (∼34 years), 1980 (∼44 years), 1990 (∼54 years) and 2001 (∼65 years) using AQI. Accumulation of air pollution exposure across the life course was conveyed as area under the curve; we first multiplied exposures with the number of years in the respective time band and then summed them (i.e. [AQI 1935 × 7] + [AQI 1950 × 17] + [AQI 1970 × 16] + [AQI 1980 × 10] + [AQI 1990 × 10] + [AQI 2001 × 11]). We found stronger correlations between sensitive periods closer to each other in time, and between sensitive periods and accumulation (Figure S4). To adjust for confounding before model selection, we produced model residuals after regressing life-course models on their specific confounders picked individually using the DAG (Table S1).

In the second stage, we estimated effect sizes for selected life-course models utilising linear mixed-effects regression with random intercepts making use of all available epigenetic data across waves 1 and 4. Random slopes for chronological age were not considered as trajectories because age acceleration did not change significantly during shorter follow-ups.^22^ We included the same life-course specific confounders in the regression as used in the model selection stage. In addition to primary findings for the AQI (coefficients expressed as 1 standard deviation [SD] increase), we also reported results separately for PM_2.5_, SO_2_, NO_2_, and O_3_ (expressed as 1-μ*g/m^3^*) with FDR adjustment for *p*-values. Moreover, to aid the comparison of the magnitude of associations across all findings, we presented fully standardized coefficients (i.e. both exposure and outcome) as βs. Potential sex-differences identified in the male and female samples were formally investigated in the non-stratified sample using the interaction term of AQI × sex.

Five sets of analyses were carried out to assess the sensitivity and robustness of our findings. First, we adjusted models for white blood cell counts as fixed effects (i.e. CD8T, CD4T, NK, Bcell, Mono, Gran), extracted from the same blood sample as for DNAm using Houseman’s algorithm.^50^ Second, methylation data were generated across three separate laboratory experiments (Table S2). We reran the main models including the set of experiments (set1, set2, set3) and batch variables (i.e. position on array, array, plate, hybridisation date) as random effects. Third, instead of accounting for only relevant life-course confounders in the model selection stage, we regressed life-course variables on all covariates to reduce the likelihood of unmeasured confounding (e.g. there is high correlation between BMI assessments from adolescence onwards,^51^ but LBC1936 only captures BMI data from late adulthood). After selecting the best-fit life-course models, we also presented main models with adjustments for all confounders. Fourth, we ran two-pollutant models to explore co-pollutant confounding whereby each pollutant was added as a covariate in the models for every other pollutant. Last, air pollution exposure around 1935 was originally derived from addresses between 1936-1942; in a *post-hoc* analysis we present findings for relevant models using 1936 addresses only, providing a stronger case for *in utero* exposure.

All analyses were conducted in R 4.1.0.^52^

## RESULTS

### Study population

The sample included 525 individuals; 437 participated in wave 1, 489 in wave 2, 455 in wave 3 and 401 in wave 4. The majority of excluded LBC1936 participants dropped out before the lifegrid questionnaire was distributed in 2014 (*n*=387) or did not provide address history (*n*=111). A comparably smaller number of individuals had missing information on air pollution exposure in at least one time period, due to living outside of the UK (*n*=22), or missing covariate or outcome data (*n*=46) (Figure 1). Excluded individuals smoked more frequently in childhood and adulthood, had higher BMI at age 70 and achieved a lower OSC in adulthood (Table 1). Participants’ residential exposure to air pollution changed markedly during their life course, with PM_2.5_ and SO_2_ levels monotonically dropping from the 1950s, whereas exposure to NO_2_ and O_3_ increased across the life course (Figure 2; Table S3). Between waves 1 and 4, participants aged on average 10 years; the biological ageing process during these years materialised in increasing epigenetic clock estimates and shortening DNAmTL (Table S4).

**Figure 1.**
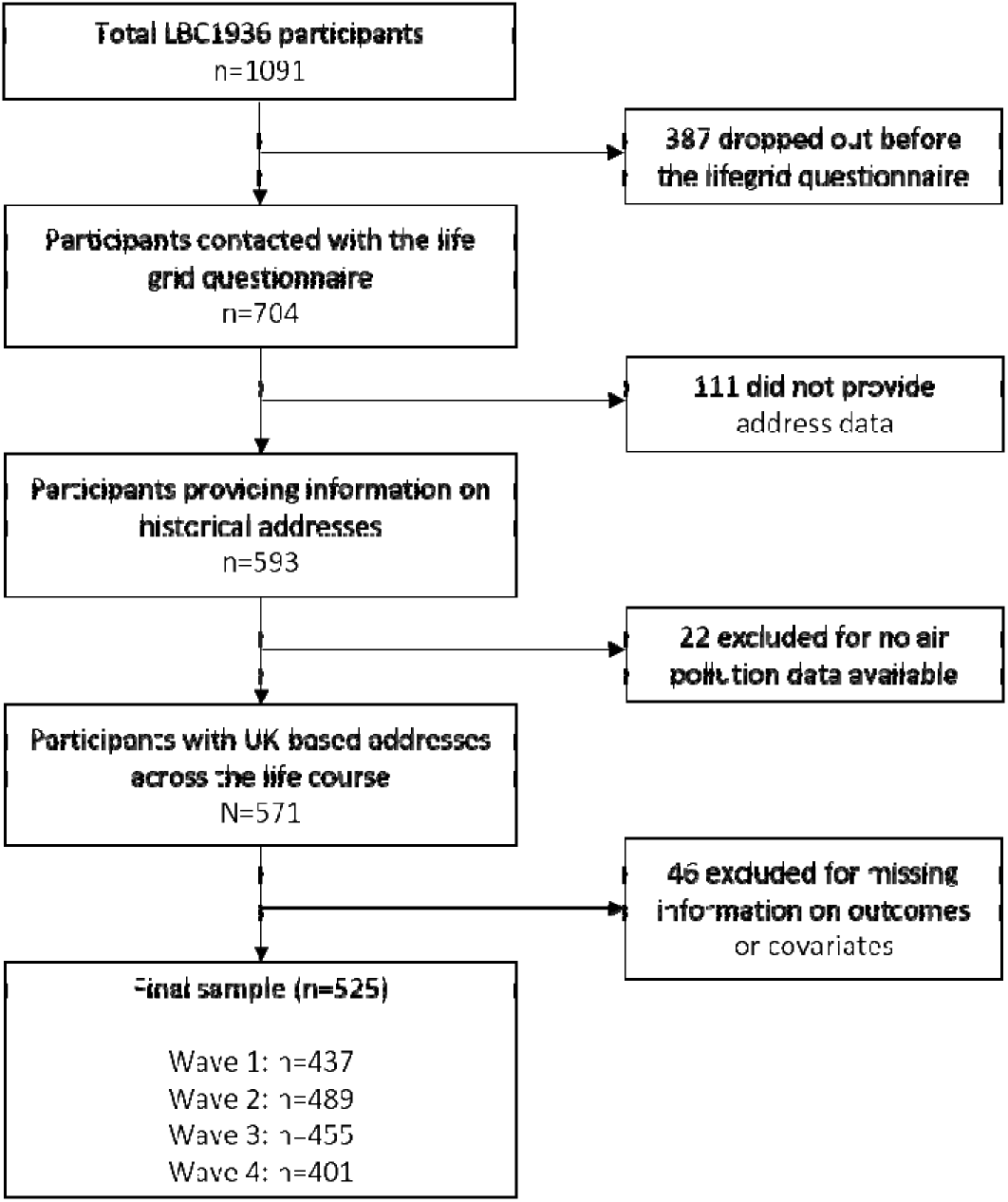
Flowchart indicating sample selection. We utilised wave 2 data for model selection, the complete sample with repeated measurements for model estimation.

**Figure 2.**
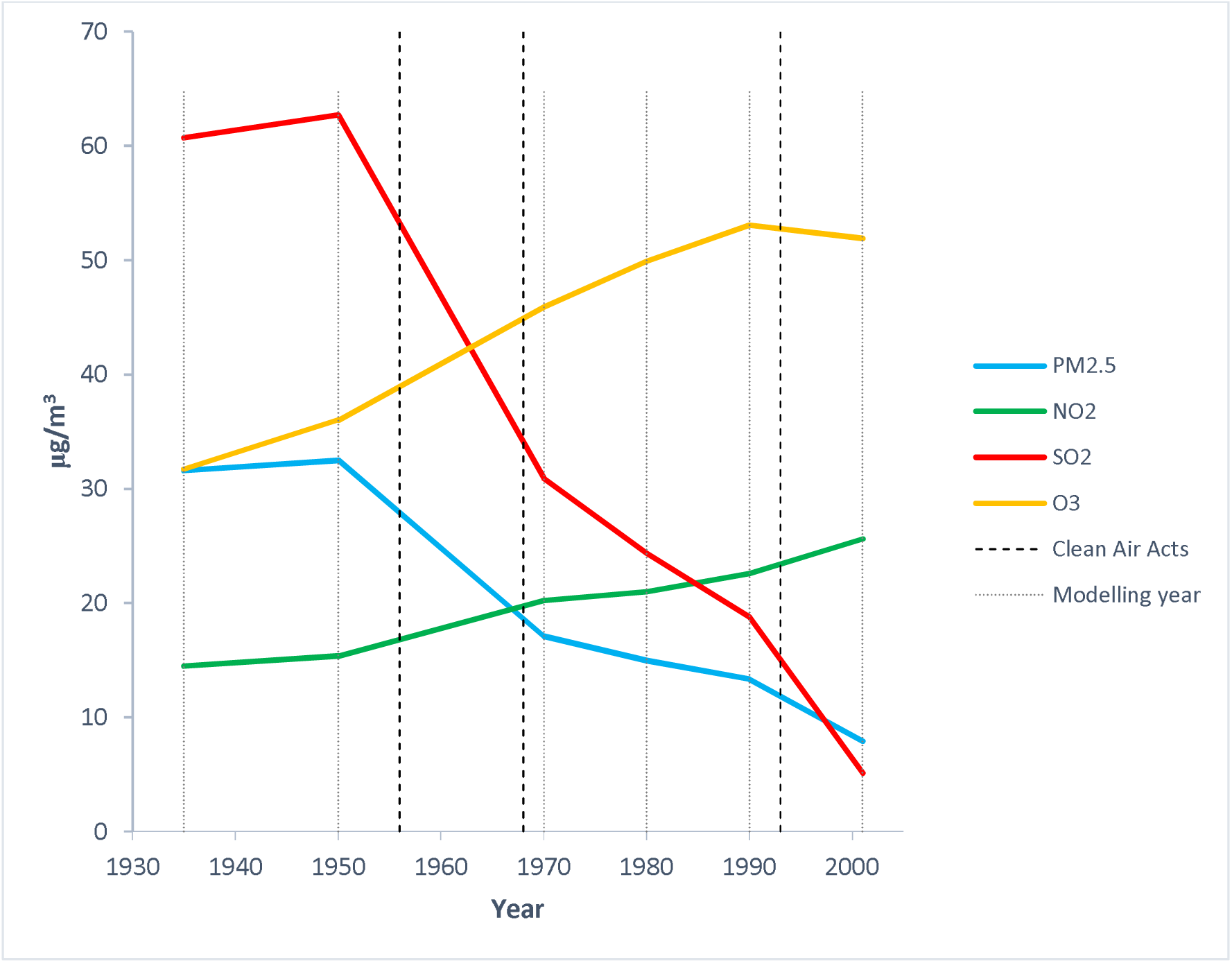
Annual average exposure to ambient air pollution among LBC1936 participants (*n*=525). Concentrations were estimated using the EMEP4UK atmospheric chemistry transport model for the modelling years of 1935, 1950, 1970, 1980, 1990, and 2001 (dotted lines). Clean Air Acts of 1956, 1968 and 1993 – key regulatory mechanisms reducing the air pollution during the study period in the United Kingdom – are signalised as dashed lines. Recommended WHO air quality guideline levels (2021) are 5 *µg/m^3^* for annual PM_2.5_, 10 *µg/m^3^* for annual NO_2_ exposure. For SO_2_ and O_3_, shorter averaging times are available with the 24-hour level of *40 µg/m^3^* for SO_2_, and the peak season level of *60 µg/m^3^* for O_3_.

**Table 1.**
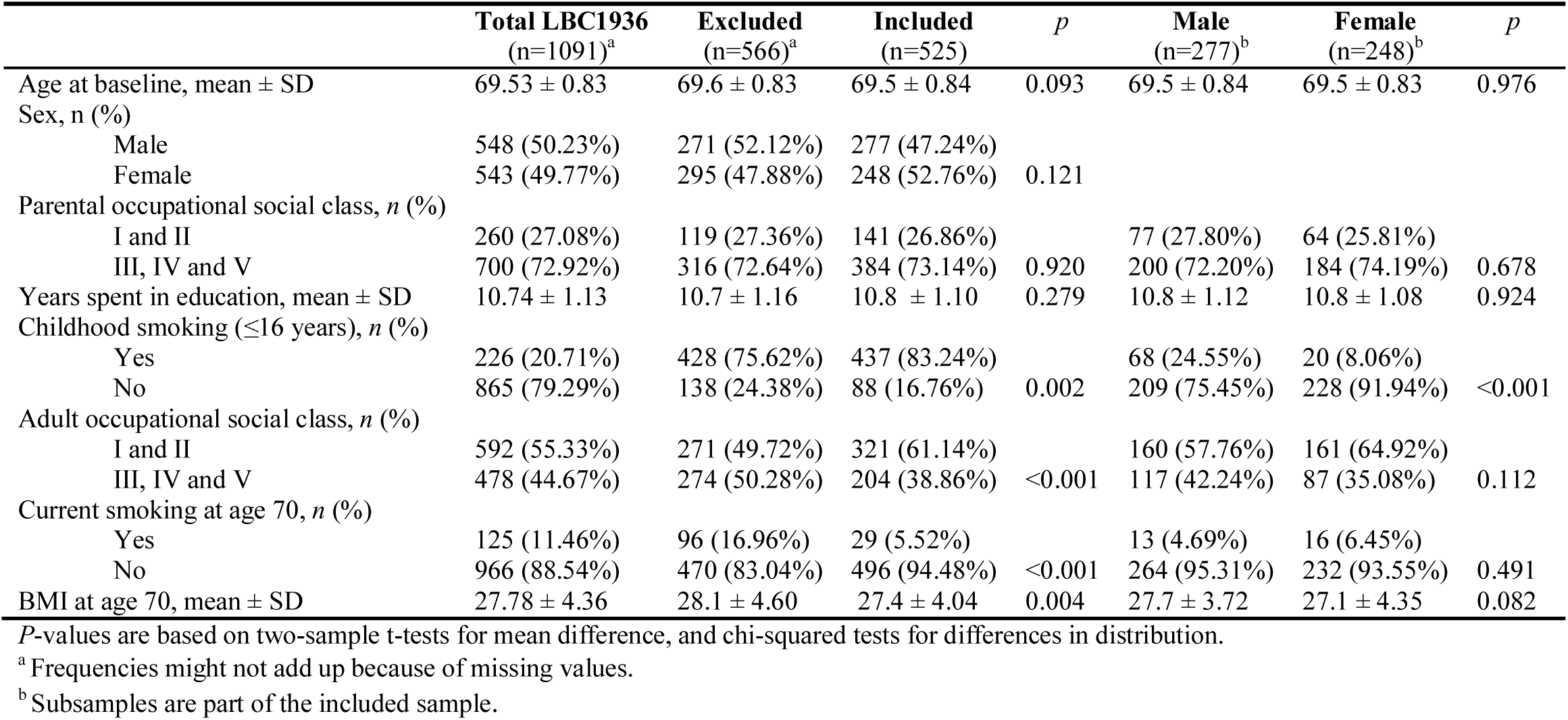
Descriptive statistics for included and excluded participants, Lothian Birth Cohort 1936

### Stage 1: Identifying best-fit life-course models

For the model selection stage, we utilised outcome data from wave 2 as providing the largest sample size (*n*=489), and thus allowing the best approximation of the total sample. The covariance test for the lasso indicated that air pollution exposure around 1970 significantly reduced outcome variance for Horvath DNAmAge (*p*=0.039; *p_FDR_*=0.193), accounting for 0.9% of the residual variance after adjusting for life-course confounders selected based on the DAG (i.e. age, sex, parental OSC, childhood smoking, years spent in education). In the non-stratified sample, air pollution was not associated with any other biomarkers (Table 2).

**Table 2.** Selecting best-fit life-course models for the association between Air Quality Index and markers of biological ageing in the Lothian Birth Cohort 1936.

| Outcome | Total<br>(n=489) |  |  |  | Male<br>(n=265) |  |  |  | Female<br>(n=224) |  |  |  |
| --- | --- | --- | --- | --- | --- | --- | --- | --- | --- | --- | --- | --- |
|  | Model | R<sup>2</sup> | p | p<sub>FDR</sub> | Model | R<sup>2</sup> | p | p<sub>FDR</sub> | Model | R<sup>2</sup> | p | p<sub>FDR</sub> |
| Horvath DNAmAge | SP 1970 | 0.009 | 0.039 | 0.193 | SP 1970 | 0.007 | 0.286 | 0.715 | SP 1980 | 0.005 | 0.521 | 0.869 |
| Hannum DNAmAge | SP 1950 | 0.004 | 0.259 | 0.586 | SP 1950 | 0.005 | 0.527 | 0.752 | SP 1990 | <0.001 | 0.990 | 0.991 |
| DNAm GrimAge | SP 1935 | 0.001 | 0.830 | 0.432 | SP 1950 | 0.001 | 0.792 | 0.792 | SP 1935 | 0.002 | 0.778 | 0.973 |
| DNAm PhenoAge | SP 1980 | 0.008 | 0.083 | 0.210 | SP 1980 | 0.003 | 0.602 | 0.752 | SP 1980 | 0.009 | 0.295 | 0.737 |
| DNAmTL | SP 1980 | 0.001 | 0.841 | 0.841 | SP 1980 | 0.031 | 0.003 | 0.014 | SP 1935 | 0.026 | 0.008 | 0.042 |

Life-course models were progressively adjusted for confounders: SP 1935 for age, (sex,) parental occupational social class; SP 1950 additionally for childhood smoking; SP 1970 and SP 1980 additionally for years spent in education; SP 1990 additionally for adult occupational social class and BMI at age 70. We provide false discovery rate (FDR) adjusted p-values. DNAmTL – DNAm telomere length; SP – sensitive period.

Among males (*n*=265), we found that a sensitive period around 1980 was the most appropriate life-course model, accounting for 3.1% of the residual variance in DNAmTL (*p*=0.003; *p_FDR_*=0.014). In the female subsample (*n*=224), air pollution exposure around 1935 explained 2.6% of the residual variance in DNAmTL (*p*=0.008; *p_FDR_*=0.042) (Table 2).

Although the selected life-course model in the non-stratified sample (i.e. air pollution exposure around 1970 and Horvath DNAmAge) did not pass adjustment for false discovery rate, we carried forward all three models to the model estimation stage to test findings in the full sample.

### Stage 2: Estimating best-fit life-course models

Selected life-course models (i. air pollution around 1970 for Horvath DNAmAge; ii. air pollution around 1980 for DNAmTL among male; and iii. air pollution around 1935 for DNAmTL among female) were estimated in mixed-effects regressions using all available epigenetic data (*n*=525; *obs*=1782) and adjusting for life-course specific confounders.

We found that 1 SD increase in the composite AQI around 1970 was associated with a 0.622 year (95% CI: 0.151, 1.094; *p*=0.010) higher epigenetic age measured in Horvath DNAmAge (Table 3); no sex differences were identified (*b*=0.397, 95% CI: -0.551, 1.346; *p*=0.412; Figure S5). Estimating the associations for single air pollutants showed that 1 μ*g/m^3^* increase in PM_2.5_ and SO_2_ levels in 1970 was associated with an epigenetic age increase of 0.317 (95% CI: 0.024, 0.578; *p*=0.034) and 0.087 (95% CI: 0.007, 0.167; *p*=0.034) years, respectively. Although just above threshold-level, none of the findings for Horvath DNAmAge passed FDR adjustment (Table 3).

**Table 3.** Exposure to air pollution and accelerated ageing in the Lothian Birth Cohort 1936

| Exposure | Total<br>(*n*=525, *obs*=1782)<br>Horvath DNAmAge<sup>a</sup><br>Air pollution in 1970 |  |  | Male<br>(*n*=277, *obs*=950)<br>DNAmTL<sup>b</sup><br>Air pollution in 1980 |  |  | Female<br>(*n*=248, *obs*=832)<br>DNAmTL<sup>c</sup><br>Air pollution in 1935 |  |  |
| --- | --- | --- | --- | --- | --- | --- | --- | --- | --- |
|  | Estimate (95% CI) | *p* | *p<sub>FDR</sub>* | Estimate (95% CI) | *p* | *p<sub>FDR</sub>* | Estimate (95% CI) | *p* | *p<sub>FDR</sub>* |
| Air Quality Index (in 1 *SD*) | 0.622 (0.151, 1.094) | 0.010 | 0.050 | -0.035 (-0.057, -0.014) | 0.002 | 0.005 | -0.036 (-0.059, -0.013) | 0.002 | 0.005 |
| Air pollutants (in 1 $\mu\text{g}/\text{m}^3$) | | | | | | | | | |
| □PM<sub>2.5</sub> | 0.301 (0.024, 0.578) | 0.034 | 0.057 | -0.023 (-0.038, -0.008) | 0.003 | 0.005 | -0.003 (-0.004, -0.001) | 0.003 | 0.005 |
| □SO<sub>2</sub> | 0.087 (0.007, 0.167) | 0.034 | 0.057 | -0.005 (-0.009, -0.001) | 0.025 | 0.031 | -0.001 (-0.002, -0.000) | 0.003 | 0.005 |
| □NO<sub>2</sub> | 0.111 (-0.003, 0.226) | 0.057 | 0.071 | -0.007 (-0.012, -0.003) | 0.003 | 0.005 | -0.006 (-0.011, -0.002) | 0.006 | 0.008 |
| □O<sub>3</sub> | -0.089 (-0.228, 0.050) | 0.210 | 0.210 | 0.006 (0.000, 0.012) | 0.035 | 0.035 | 0.008 (0.002, 0.015) | 0.013 | 0.013 |

Models were fitted with mixed-effects regression with random intercepts for study participants. Coefficients for the Air Quality Index are standardized, for air pollutants coefficients are expressed in $\mu\text{g}/\text{m}^3$ . We provide also false discovery rate (FDR) adjusted p-values. Abbreviations: DNAmTL – DNAm telomere length; SD – standard deviation.
<sup>a</sup>Models were adjusted for sex, age, parental occupational social class, childhood smoking and years spent in education;
<sup>b</sup>Models were adjusted for age, parental occupational social class, childhood smoking and years spent in education;
<sup>c</sup>Models were adjusted for age and parental occupational social class.

Among males (*n*=277; *obs*=950), 1 *SD* increase in the composite AQI around 1980 was associated with 0.035 kilobase (95% CI -0.057, -0.014; *p*=0.002) reduction in DNAmTL. Significant associations could be observed across all pollutants around 1980, with higher exposure of PM_2.5_ (*p*=0.003), SO_2_ (*p*=0.025) and NO_2_ (*p*=0.003) being associated with shorter, while higher exposure to O_3_ with longer, telomere length (*p*=0.035) (Table 3). In the female subsample (*n*=248; *obs*=832), we found that 1 *SD* increase in AQI around 1935 was associated with a telomere attrition of 0.036 kilobase (95% CI -0.059, -0.013; *p*=0.002); higher PM_2.5_ (*p*=0.003), SO_2_ (*p*=0.003) and NO_2_ (*p*=0.006) exposure with shorter, and higher O_3_ (*p*=0.013) with longer telomeres. All findings reported for DNAmTL passed FDR adjustment (Table 3). Testing sex-differences confirmed stronger DNAmTL attrition among males with higher air pollution exposure around 1980 (AQI: *b*=-0.042, 95% CI: -0.074, - 0.010, *p*=0.010); sex-interaction for air pollution around 1935 and DNAmTL was above threshold level (*b*=0.032, 95% CI: -0.000, 0.064; *p*=0.051) (Figure S5).

Finally, fully standardized coefficients are presented in Figure 3. This indicates that all associations were of small effect size.^53^ Associations with DNAmTL were generally numerically stronger than for Horvath DNAmAge, and coefficients for O_3_ were weaker compared to PM_2.5_, SO_2_ and NO_2_. (However, these were not formally tested, and we point out that the 95% CIs overlap in all cases.)

**Figure 3.**
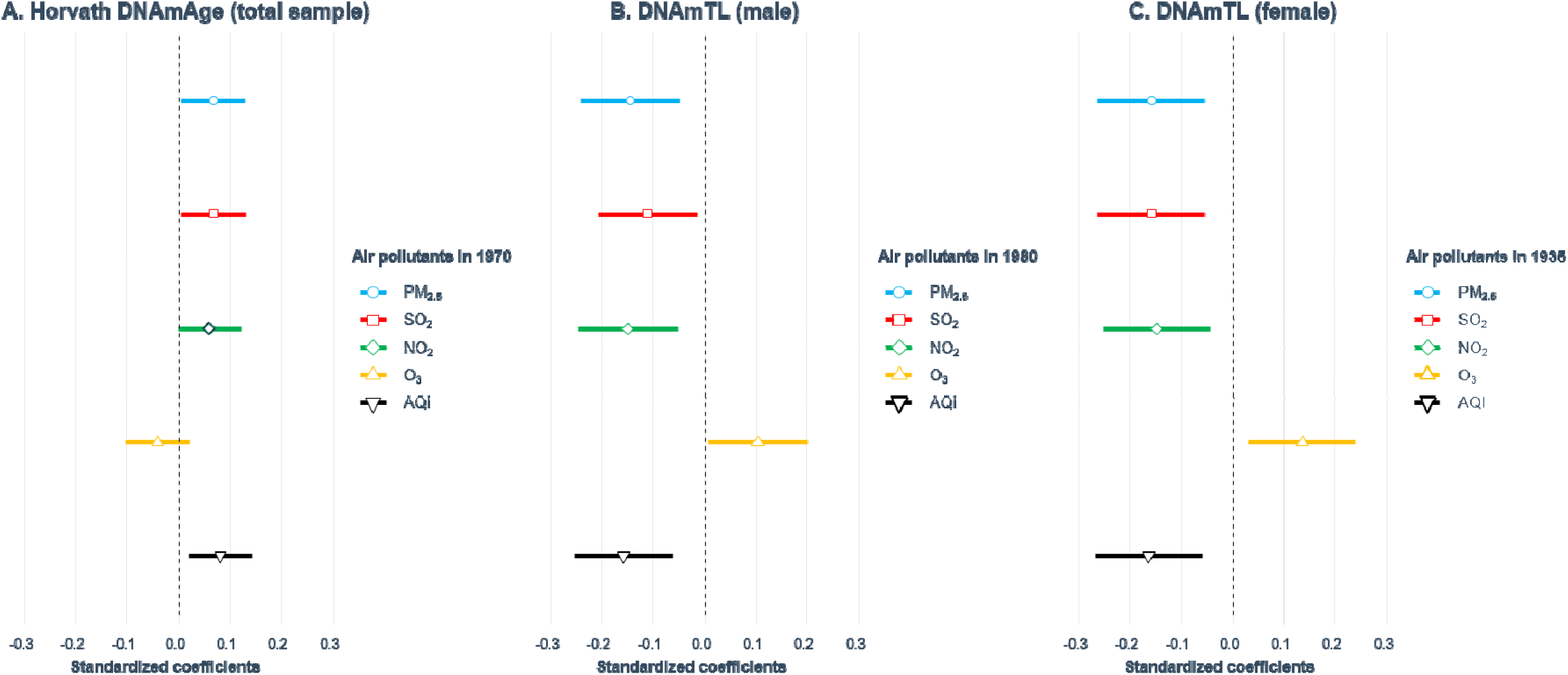
Association between air pollution and DNAm-based biomarkers in the Lothian Birth Cohort 1936 (*n*=525). Fully standardized coefficients (β) and their 95% CIs are presented to aid comparison of effect sizes. Model A was adjusted for sex, age, parental occupational social class, childhood smoking and years spent in education; Model B for age, parental occupational social class, childhood smoking and years spent in education; and Model C for age and parental occupational social class. Abbreviations: AQI – Air quality Index; DNAmTL – DNAm telomere length.

### Sensitivity and robustness analyses

Adjusting for white blood cell counts as fixed effects (Table S5), or experiment variables as random effects (Table S6) did not substantially change the associations between air pollution and DNAm-based biomarkers. Rerunning LARS model selection procedure after adjusting life-course models with all confounders independently of their timing during the life course (i.e. sex, age, parental OSC, childhood smoking, years spent in full-time education, adult OSC, smoking and BMI at age 70) resulted in the same selection of the three life-course models before FDR adjustment (Table S7). Estimating models after these adjustments did not alter the results (Table S8).

In two-pollutant models, we did not find evidence for one pollutant having an independent effect above any other (Table S9). Rather the single pollutant models with comparable effect sizes and the two-pollutant models with high collinearity indicated that the generalised effect of air pollution cannot be attributed to any single component of poor air quality in the current sample. Still, we observed that around 1935 and 1970 PM_2.5_ and SO_2_, around 1980 PM_2.5_ and NO_2_ remained more often under *p*<0.1 after adjusting for co-pollutants; in comparison to its co-pollutants, O_3_ was weaker (and never significantly) associated with biological ageing markers. Finally, when we matched 1935 air pollution estimates with 1936 addresses only (instead of addresses between 1936 and 1942), the magnitude of association between the revised AQI in 1935 and DNAmTL increased by approximately 25% to -0.045 (95% CI - 0.068, -0.022; *p*<0.001; *n*=246), highlighting the importance of *in utero* exposure.

## DISCUSSION

Our study based on 525 older Scottish adults examined the relationship between life-course air pollution exposure and DNAm-based biomarkers. Out of a large number of tested associations, our analyses identified three significant ones, reporting a link between greater exposure to air pollution during the first half of life and older-appearing markers of biological ageing in later life. Exposure to air pollution around the age ∼34 (i.e. 1970) was associated with epigenetic age acceleration measured with Horvath’s epigenetic clock. Shorter telomere lengths estimated from DNAm were evident among males with higher exposure to air pollution at age ∼44 (i.e. 1980) and among females with higher *in utero* air pollution exposure (i.e. 1935). Effect estimates reported across this study were small in magnitude and for Horvath’s epigenetic clock did not pass FDR adjustment. Various sensitivity and robustness checks confirmed our findings.

Associations between exposure to air pollution, including PM_2.5_, and DNAm among various age groups have been reported previously (see^25, 54^ for reviews). However, those studies focussing on epigenetic clocks mainly utilised samples capturing middle-aged and older adults alongside contemporaneous exposure measures. An early investigation utilising the Normative Ageing Study in the United States, an all-male cohort of 589 individuals in their 70s, found that average residential PM_2.5_ exposure 365 days prior to outcome assessment was significantly associated with age acceleration based on Horvath DNAmAge.^23^ Further analyses suggested that CpGs contributing to this association were mapping genes involved in lung pathologies^23^ and, among the heterogeneous chemical components of PM_2.5_, sulfate and ammonium were most associated with accelerated ageing.^55^ Other studies using the same dataset did not find an association between long-term PM_2.5_ exposure and Hannum DNAmAge^55^ or DNAm PhenoAge^56^; but two chemical components of PM_2.5_ (i.e. lead and calcium) led to accelerated ageing measured with DNAm PhenoAge.^56^ The Cooperative Health Research in the Region of Augsburg (KORA) study in Germany with 1777 older participants confirmed the link between higher annual PM_2.5_ exposure at the time of data collection and extrinsic epigenetic age acceleration derived from Horvath’s epigenetic clock (i.e. regressing the clock measure on blood immune cell counts and age); however, associations (and their direction) differed between males and females.^28^ More recently, findings based on 2747 women aged 35-74 in the Sister Study from the United States indicated age acceleration using Hannum DNAmAge when exposed to higher NO_2_ levels (estimated as average 1-year exposure prior to study enrolment), while only clusters of PM_2.5_ components were associated with Horvath DNAmAge and DNAm PhenoAge.^27^ Whereas these previous reports described the associations between air pollution exposure and faster epigenetic clocks in different samples, none has been able to look at how exposure at different epochs relates to DNAm in older age. Our study not only confirms the relationship between accelerated epigenetic ageing based on Horvath DNAmAge among individuals exposed to higher air pollution,^23, 28^ but also extends the literature suggesting a sensitive period around the age of 34. Still, we would highlight that this finding did not survive correction for multiple testing, introducing uncertainty in the interpretation.

We observed significantly shorter DNAmTL among older males exposed to higher air pollution levels around 1980. These findings corroborate and extend prior work relating to leukocyte telomere length. A systematic review among adults found that long-term exposure to air pollution is associated with shorter telomere length, whereby pollution likely increases the replication rate of cells and telomere loss during cell replication.^9^ Findings from the KORA study indicated sex-differences in the air pollution (i.e. black carbon) and telomere length relationship, with significant attrition found only among males.^28^ DNAmTL is a robust measure of telomere-associated ageing, which is related to cell replication and cellular ageing, distinct from epigenetic ageing.^43^ CpGs used to derived epigenetic clocks and DNAmTL are not overlapping; CpGs for DNAmTL are located near cadherin and cell signalling genes.^43^ DNAmTL has been shown to outperform leukocyte telomere length in predicting health-related outcomes^43^; and, to our knowledge, it has not been used before to explore the air pollution-biological ageing relationship.

Another key finding of this study showed that *in utero* exposure to air pollution was associated with shorter DNAm-based telomere length among females. This is supported by a recent systematic review concluding that prenatal exposure to air pollution is linked to global and specific alterations in DNAm levels and to telomere attrition, whereby the beginning of the pregnancy is a potentially susceptible period.^24^ Particles can translocate into or across the placenta and induce oxidative stress; production of reactive oxygen species leads to DNA damage and epigenetic alterations.^57^ In turn, changes in methylation levels in the embryonic development are associated with abnormal development.^58^ Mediation analyses have confirmed the role of DNAm in the pathway between air pollution exposure and foetal growth,^59^ and later life health outcomes.^60^ In our study, *in utero* exposure was associated with DNAmTL only among females, which is not unexpected given sex-differences in the impact of air pollution on pregnancy outcome.^61^ In line with our finding, a study from Mexico found shorter leukocyte telomere length among female but not male newborns after higher maternal PM_2.5_ exposure.^62^ Similarly, prenatal exposure to organic pollutants in the Shanghai Allergy Cohort was linked to shorter leukocyte telomere length at birth among females, and mediation analyses highlighted the role of elevated oxidative stress.^63^

The air we breathe typically contains a mixture of multiple pollutants. Although we provided estimates for PM_2.5_, NO_2_, SO_2_ and O_3_ separately, these were highly correlated making it impossible to properly disentangle their effects on DNAm-based biomarkers, which is generally challenging in the absence of experimental data. NO_2_ and SO_2_ for example are emitted as primary pollutants, whereas PM_2.5_ is a mixture of primary and other secondary pollutants such as ammonium nitrate and ammonium sulphate; fossil fuel combustion sources contribute the major share of all three pollutants. O_3_ is not directly emitted into the air, but it is produced through complex chemical reactions in the atmosphere driven by NO_2_ and Volatile Organic Compounds.^1^ Due to the titration effect, O_3_ is depleted in areas where high NO_x_ emissions are present, which results to O_3_ being negatively correlated with NO_2_ (and to a smaller degree with PM_2.5_ and SO_2_). This might be particularly apparent when concentrations are aggregated at higher units. While there is some evidence of a positive association between O_3_ exposure and leukocyte telomere length among critically ill patients,^64^ our unexpected findings on O_3_ and slower biological ageing might be an artefact given the above presented correlation pattern between pollutants. Two-pollutant models further suggested that overall results were mainly driven by PM_2.5_, NO_2_, and SO_2_, making a plausible positive association between O_3_ and DNAmTL less likely.

### Strengths and limitations

Analyses in this study were based on over 500 individuals, 1700 epigenetic samples with 5 different DNAm-based biomarkers measured in older age, and on pollution exposure estimated across the lifecourse.^31^ We utilised robust and validated methods of measuring different aspects of biological age acceleration; it is particularly important for telomere-related ageing, where the traditional methods of estimating leukocyte telomere length can be challenging.^43^ A unique feature of the cohort is the presence of life-course addresses,^29^ which made it possible to link individual residence to historical air pollution concentrations. Whereas retrospective recall is well-known to be prone to bias, the lifegrid approach with lightbulb prompt indicates that residential address recall among older people shows good accuracy.^32^ We are unaware of any other datasets that can allow interrogation of these relationships in the same individuals with coverage from *in utero* to the 8^th^ decade of life. Our analytical approach was particularly useful when competing life-course hypotheses were equally plausible^46^ and we were able to test these without over-inflating coefficients and biasing the hypothesis test.^49^

Still, several limitations need to be considered that potentially affect the interpretation of the results. First, LBC1936 compiles an ethnically homogenous sample of Scottish adults, thereby limiting the generalizability of our findings. Moreover, address data were collected when participants were in their late 70s, leaving healthier individuals in our analytical sample, introducing not only selection but also survival bias.^31^ Second, residential address in 1936 indicated by the participants may not correspond with pre-birth location, leading to exposure misclassification. Third, the selection of the best-fit life-course models was based on 489 out of 525 participants, as DNAm data were not available for the complete sample in any of the follow-up waves. Given that age acceleration trajectories are relatively stable in our cohort,^22^ we do not assume that conducting model selection with waves other than wave 2 would have altered our findings. However, unavailability of data for some participants might have reduced statistical power to identify all relevant life-course models. Fourth, there is a large degree of uncertainty when estimating historical concentrations of air pollution. Previous research using the same atmospheric chemistry transport modelling as our study has presented the longer-term public health improvements due to air pollution control measures set up.^38^ However, the further back in time emissions of relevant air pollutants have to be estimated, the larger uncertainties are with regard to their spatial distribution, as well as their volume. In addition, while atmospheric chemistry transport models are routinely and widely evaluated against observations in present-day conditions,^36^ there are few, if any, reliable observations available for such validations prior to the 1970s (for some pollutants not before the late 1980s). Also, atmospheric chemistry transport models rely on meteorological driver data (in the case of the EMEP4UK model, the Weather Research Forecast [WRF] 3.7.1)^65^ to represent atmospheric transport and chemical transformation processes. The WRF model included data assimilation (Newtonian nudging) of the numerical weather prediction model meteorological reanalysis from the US National Centers for Environmental Prediction/National Center for Atmospheric Research Global Forecast System at 1 degree resolution, every 6 hours.^66^ While these aspects clearly contribute to uncertainties in the estimates in ambient air pollutant concentrations and thus exposures, the consistent model setup and handling of input data (i.e. anthropogenic emissions and meteorological drivers) means that relative changes in the spatial distribution of concentrations can be considered to be fairly robust. Fifth, the spatial resolution of ∼5×6 km^2^ used in this study is likely too coarse, especially for urban areas, and may lead to underestimating exposures and health impacts.^67^ This is pertinent for the 2001 air pollution exposure estimate: while during their life course LBC1936 participants resided in various places across the UK, they all lived around Edinburgh when the cohort started, reducing the heterogeneity of exposure. Concentration of air pollutants were in excess of the WHO air quality guidelines^1^ during much of the study period; still, we cannot compare findings directly with contemporaneous high-excess exposure settings (e.g. Mexico City),^62^ as the sources of emission likely differ in historical and geographical context (e.g. combustion of wood, coal or oil-based products).^6^ Finally, we were only able to use residential addresses to estimate air pollution exposure; incorporating school, work and other key locations could have led to more precise findings.

Future studies exploring the life-course association between air pollution and biological ageing should address the above limitations. On the exposure site, historical air pollution data at finer scale resolution may overcome the challenges originating from the very high correlation between pollutants and provide further heterogeneity of exposure. Telomere length based on DNAm proved a valuable biomarker in our study; future investigations should further explore its utility in understanding how environmental exposures can ‘get under the skin’. Finally, research should aim to replicate our results in larger, nationally representative cohort studies with more diverse populations.

## CONCLUSIONS

This study utilised historical air pollution concentrations of PM_2.5_, SO_2_, NO_2_ and O_3_ and applied the life-course approach for the first time to contribute to the understanding of air pollution and biological ageing. We found that exposure to lower air quality at earlier stages of the life (i.e. *in utero*, young-to-mid adulthood) can have a modest but detectable association with epigenetic and telomere-associated ageing, which likely persists across the entire life course. This study demonstrated the utility of DNAmTL in environmental research, a biomarker of cellular ageing, which seems to be particularly susceptible to air pollution exposures. Future studies should explore options to refine historical air pollution data and reinforce our findings in larger cohorts. Policy actions at national-level targeting air pollution reduction can likely have long-lasting effects on the development of future generations, especially in light of findings on *in utero* effects, and contribute to healthy population ageing.

## Data Availability

Data-availability statement: The LBCs study data have been the subject of many internal (within the University of Edinburgh) and external collaborations, which are encouraged. Those who have interests in outcomes other than cognitive domains are particularly encouraged to collaborate. Both LBC studies have clear data dictionaries which help researchers to discern whether the variables they wish to use are present; these provide a simple short title for each variable, alongside a longer, common-sense description/provenance of each variable. This information is available on the study website (https://www.ed.ac.uk/lothian-birth-cohorts) alongside comprehensive data grids listing all variables collected throughout both LBC studies and the wave at which they were introduced, an LBC Data Request Form and example Data Transfer Agreement. Initially, the Data Request Form is e-mailed to the Lothian Birth Cohorts Director Dr Simon R. Cox for approval (via a panel comprising study co-investigators). Instances where approved projects require transfer of data or materials outside the University of Edinburgh require a formal Data Transfer Agreement or Material Transfer Agreement to be established with the host institution. The process is facilitated by a full-time LBC database manager, there is no charge.

## ACKNOWLEDGEMENTS

The Lothian Birth Cohort 1936 study acknowledges the financial support of NHS Research Scotland (NRS), through Edinburgh Clinical Research Facility. We gratefully acknowledge the contributions of the LBC1936 participants and members of the LBC1936 research team who collect and manage the LBC data. We are grateful for Dr Riccardo E. Marioni for his feedback on the first draft of this paper.

IJD, DLM, NS, SR, MV, SRC and JP obtained and managed the data for the study. GB, IJD, NS, CWT, SRC and JP conceived and designed the study. GB performed the statistical analyses and led the manuscript preparations, drafting and revision. All authors participated in the interpretation of the findings, critically revised the manuscript and approved the final version.

This work was supported by the Economic and Social Research Council, UK (grant award ES/T003669/1). The LBC1936 study is supported by Age UK (Disconnected Mind project, which supported SEH), the US National Institutes of Health (R01AG054628, which supported SRC and IJD), and the University of Edinburgh). SRC was also supported by a Sir Henry Dale Fellowship jointly funded by the Wellcome Trust and the Royal Society (221890/Z/20/Z). The atmospheric chemistry transport modelling work was underpinned by the Natural Environment Research Council award number NE/R016429/1 as part of the UK-SCAPE programme delivering National Capability and a grant under the “Improving Health with Environmental Data” call from the Natural Environment Research Council, the Chief Scientist Office, and the Medical Research Council (NE/P010849/1).

The LBC1936 study was conducted according to the Declaration of Helsinki guidelines with ethical permission obtained from the Multi-Centre Research Ethics Committee for Scotland (MREC/01/0/56), Lothian Research Ethics Committee (wave 1, LREC/2003/2/29), and the Scotland A Research Ethics Committee (waves 2-4, 07/MRE00/58). Written consent was obtained from all participants.

## Data-availability statement

The LBCs’ study data have been the subject of many internal (within the University of Edinburgh) and external collaborations, which are encouraged. Those who have interests in outcomes other than cognitive domains are particularly encouraged to collaborate. Both LBC studies have clear data dictionaries which help researchers to discern whether the variables they wish to use are present; these provide a simple short title for each variable, alongside a longer, common-sense description/provenance of each variable. This information is available on the study website (https://www.ed.ac.uk/lothian-birth-cohorts) alongside comprehensive data grids listing all variables collected throughout both LBC studies and the wave at which they were introduced, an ‘LBC Data Request Form’ and example Data Transfer Agreement. Initially, the Data Request Form is e-mailed to the Lothian Birth Cohorts Director Dr Simon R. Cox for approval (via a panel comprising study co-investigators). Instances where approved projects require transfer of data or materials outside the University of Edinburgh require a formal Data Transfer Agreement or Material Transfer Agreement to be established with the host institution. The process is facilitated by a full-time LBC database manager – there is no charge.

For the purpose of open access, the author has applied a Creative Commons Attribution (CC BY) licence to any Author Accepted Manuscript version arising from this submission.

## Supplementary Material

**Supplementary Figure S1.**
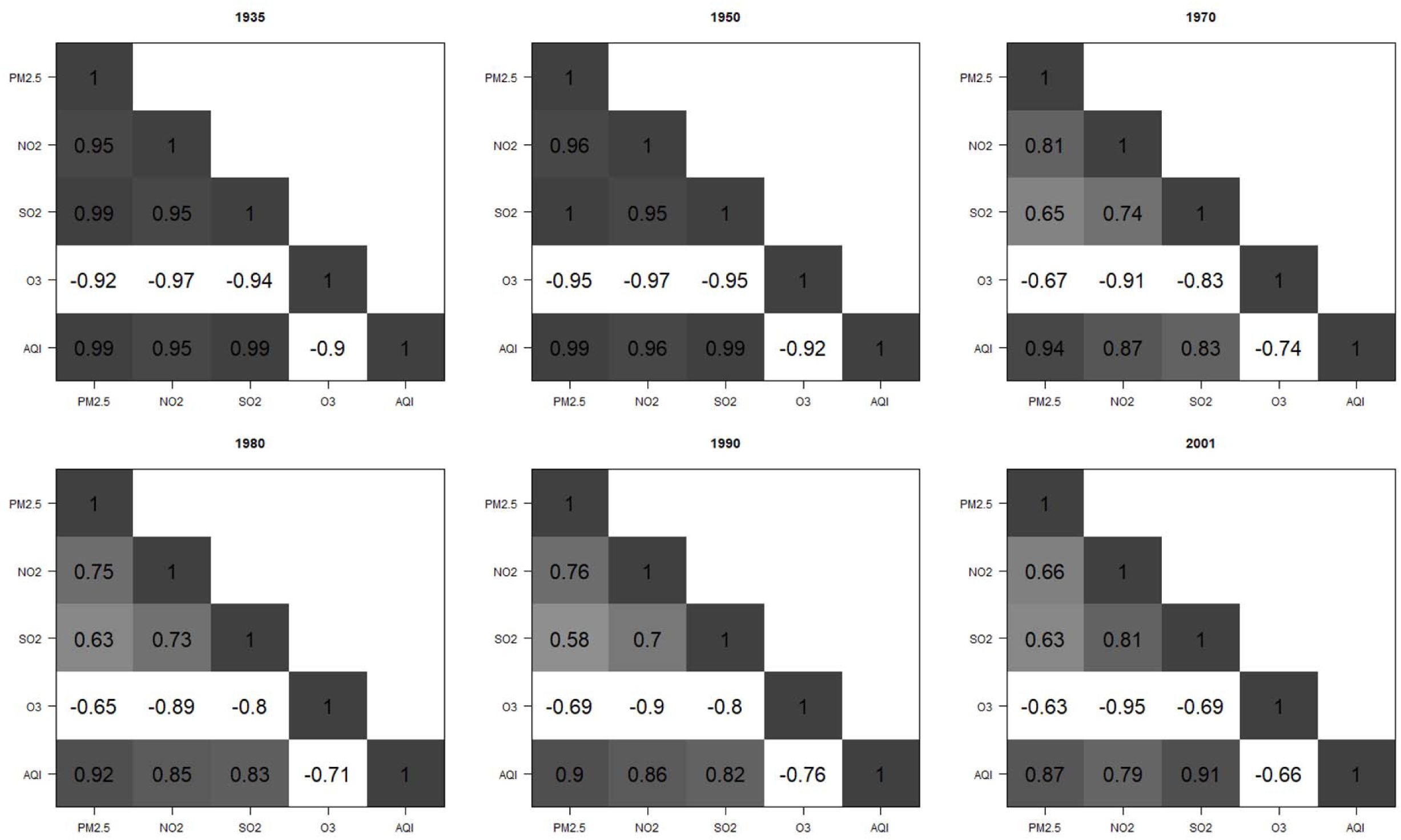
Correlation between exposure to PM_2.5_, NO_2_, SO_2_, O_3_ and Air Quality Index (AQI) across the study period. Pearson correlation coefficients are based on 525 individuals.

**Supplementary Figure S2.**
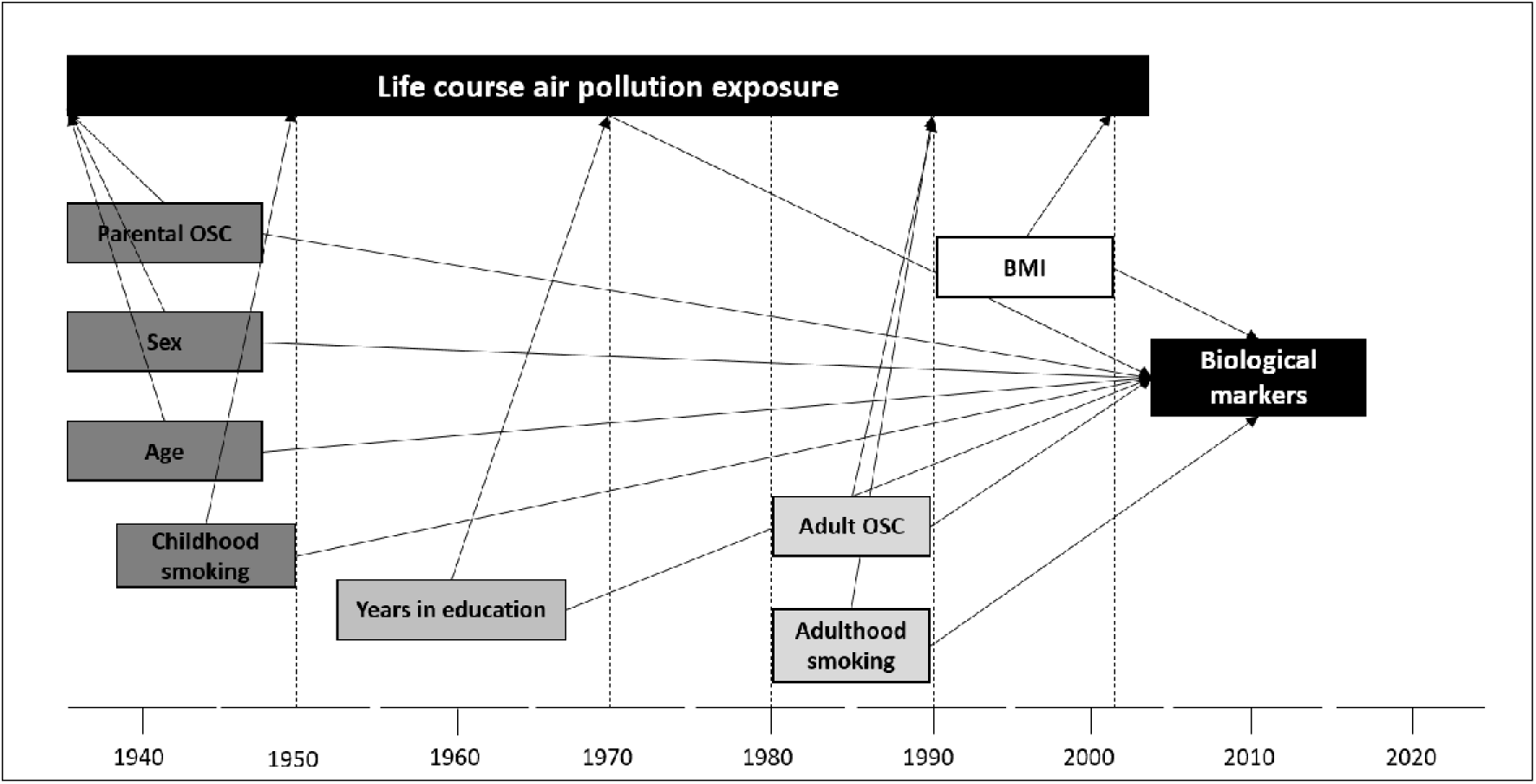
Directed acyclic graph depicting confounders of exposure to air pollution across the life course and biological ageing. Dotted lines show air pollution modelling years.

**Supplementary Figure S3.**
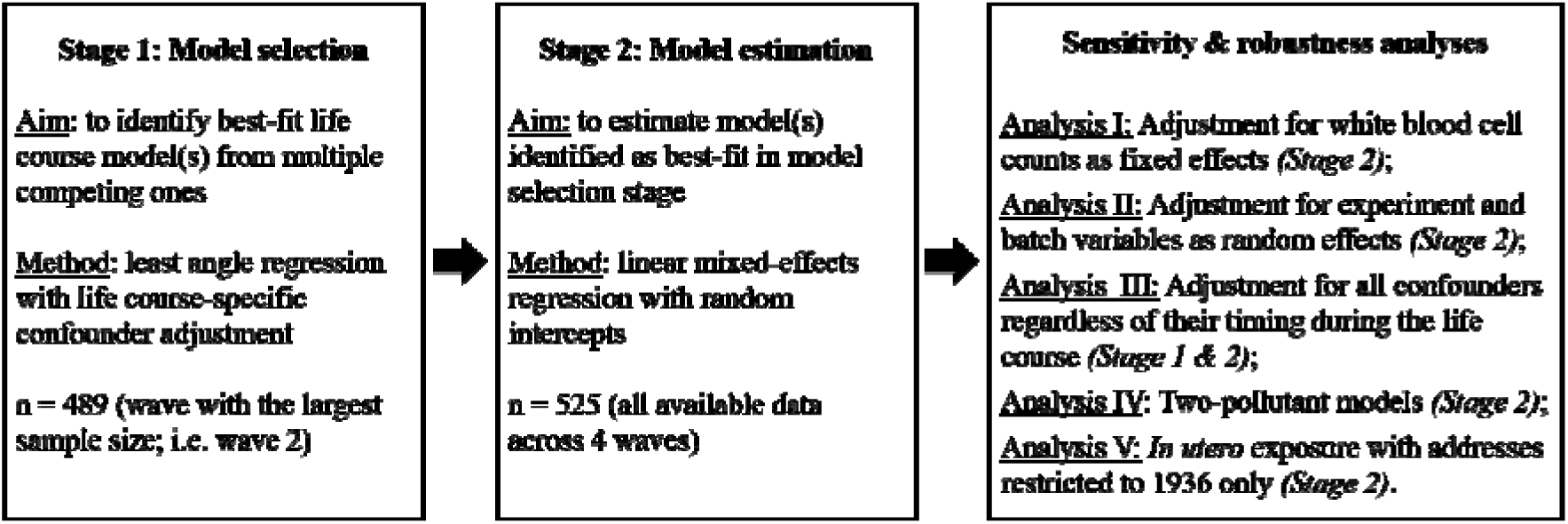
Flowchart with analyses undertaken

**Supplementary Figure S4.**
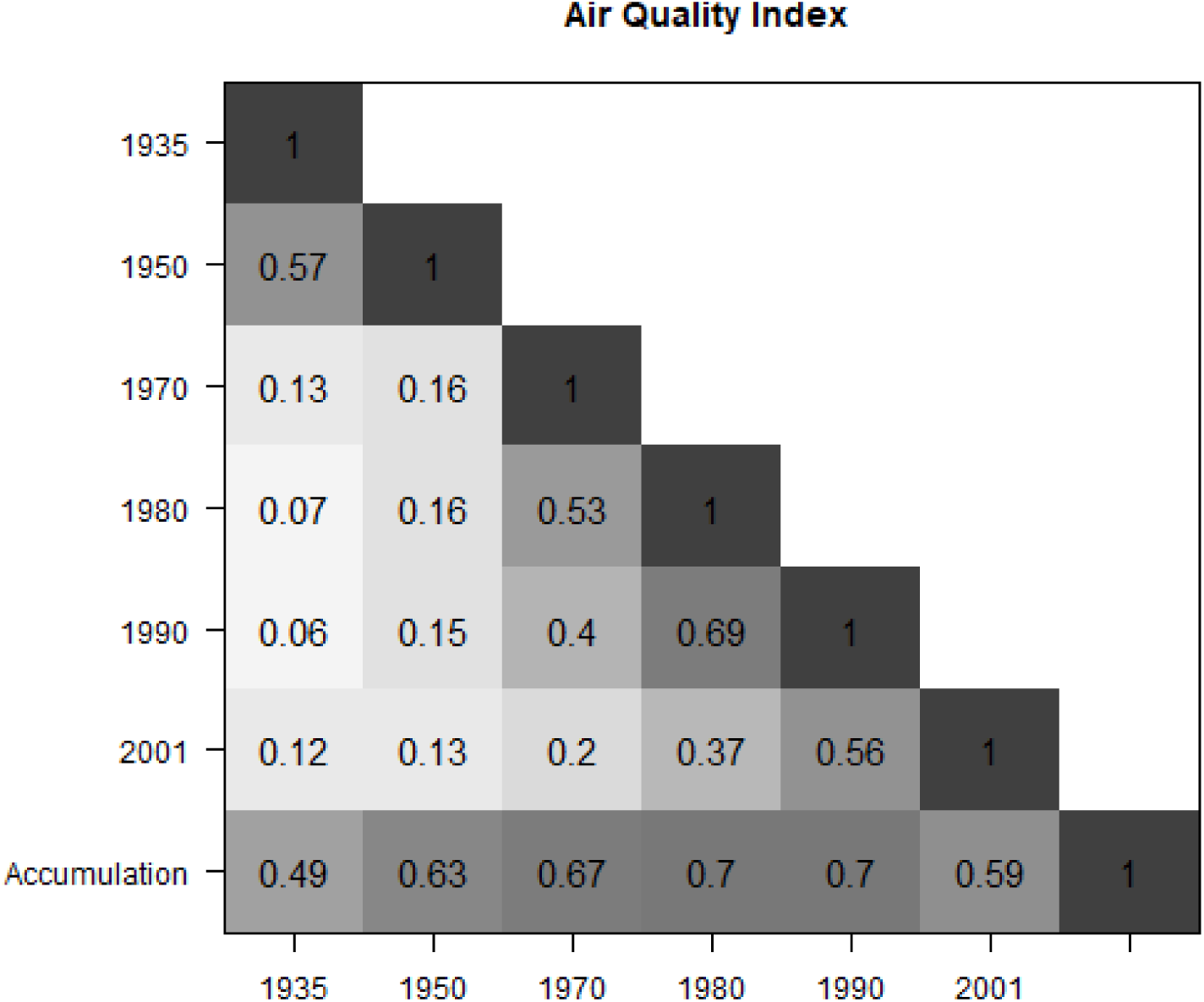
Correlation between life course models of air pollution exposure using the Air Quality Index. Pearson correlation coefficients are based on 525 individuals.

**Supplementary Table S1.** Covariate adjustment for life course models

| Life course models | Covariates |
| --- | --- |
| Sensitive period 1935 | age (+sex) + parental OSC |
| Sensitive period 1950 | age (+sex) + parental OSC + childhood smoking |
| Sensitive period 1970 | age (+sex) + parental OSC + childhood smoking + years spent in education |
| Sensitive period 1980 | age (+sex) + parental OSC + childhood smoking + years spent in education |
| Sensitive period 1990 | age (+sex) + parental OSC + childhood smoking + years spent in education + adult OSC + current smoking (measured at age 70) |
| Sensitive period 2001 | age (+sex) + parental OSC + childhood smoking + years spent in education + adult OSC + current smoking (measured at age 70) + BMI (measured at age 70) |
| Accumulation | age (+sex) + parental OSC + childhood smoking + years spent in education + adult OSC + current smoking (measured at age 70) + BMI (measured at age 70) |

Abbreviations: OSC – occupational social class.

**Supplementary Table S2.** Number of DNAm samples by set of experiments and waves

|  | Set of experiment |  |  |
| --- | --- | --- | --- |
|  | Set 1 | Set 2 | Set 3 |
| Wave 1 (*n*=437) | 422 | 15 | 0 |
| Wave 2 (*n*=489) | 207 | 277 | 5 |
| Wave 3 (*n*=455) | 186 | 269 | 0 |
| Wave 4 (*n*=401) | 0 | 0 | 401 |

**Supplementary Table S3.** Air pollution exposure in μ*g/m^3^* across the life course of LBC1936 participants.

|  | PM<sub>2.5</sub> |  | SO<sub>2</sub> |  | NO<sub>2</sub> |  | O<sub>3</sub> |  |
| --- | --- | --- | --- | --- | --- | --- | --- | --- |
| | Mean ± SD | Range | Mean ± SD | Range | Mean ± SD | Range | Mean ± SD | Range |
| 1935 | 31.59 ± 13.74 | 5.79-112.30 | 60.71 ± 31.76 | 2.86-242.72 | 14.48 ± 5.33 | 2.02-34.53 | 31.71 ± 3.67 | 17.55-41.80 |
| 1950 | 32.50 ± 12.41 | 5.96-113.26 | 62.70 ± 28.88 | 2.81-252.39 | 15.37 ± 4.94 | 1.93-37.00 | 36.03 ± 3.45 | 20.52-47.48 |
| 1970 | 17.12 ± 1.71 | 9.54-25.80 | 30.90 ± 5.97 | 5.92-54.15 | 20.22 ± 4.14 | 3.34-38.96 | 45.89 ± 3.46 | 35.94-59.87 |
| 1980 | 14.97 ± 1.43 | 7.36-22.70 | 24.36 ± 4.72 | 2.75-51.95 | 21.01 ± 4.33 | 1.82-40.71 | 49.89 ± 3.73 | 41.39-67.02 |
| 1990 | 13.35 ± 1.19 | 6.70-21.42 | 18.78 ± 3.49 | 1.36-39.97 | 22.56 ± 4.94 | 1.42-53.94 | 53.07 ± 4.09 | 35.21-74.08 |
| 2001 | 7.92 ± 0.59 | 4.79-15.95 | 5.14 ± 1.24 | 1.35-19.48 | 25.63 ± 6.53 | 3.69-76.04 | 51.90 ± 4.30 | 27.25-68.05 |

Calculations are based on 525 individuals. Abbreviations: SD – standard deviation

**Supplementary Table S4.** Markers of biological ageing among LBC1936 participants across follow-up waves

|  | Chronological age | Horvath DNAmAge | Hannum DNAmAge | DNAm GrimAge | DNAm PhenoAge | DNAmTL |
| --- | --- | --- | --- | --- | --- | --- |
| | Mean ± SD | Mean ± SD | Mean ± SD | Mean ± SD | Mean ± SD | Mean ± SD |
| Wave 1 (n=437) | 69.5 ± 0.84 | 64.8 ± 7.21 | 71.4 ± 5.67 | 66.6 ± 4.70 | 66.3 ± 7.17 | 6.75 ± 0.22 |
| Wave 2 (n=489) | 72.5 ± 0.71 | 68.1 ± 6.69 | 73.0 ± 5.77 | 69.6 ± 4.64 | 66.8 ± 7.09 | 6.71 ± 0.21 |
| Wave 3 (n=455) | 76.2 ± 0.69 | 72.1 ± 6.51 | 77.6 ± 5.74 | 72.6 ± 4.91 | 70.4 ± 7.78 | 6.62 ± 0.22 |
| Wave 4 (n=401) | 79.3 ± 0.62 | 74.8 ± 5.95 | 82.4 ± 5.55 | 75.4 ± 4.56 | 73.4 ± 6.62 | 6.55 ± 0.22 |

**Supplementary Figure S5.**
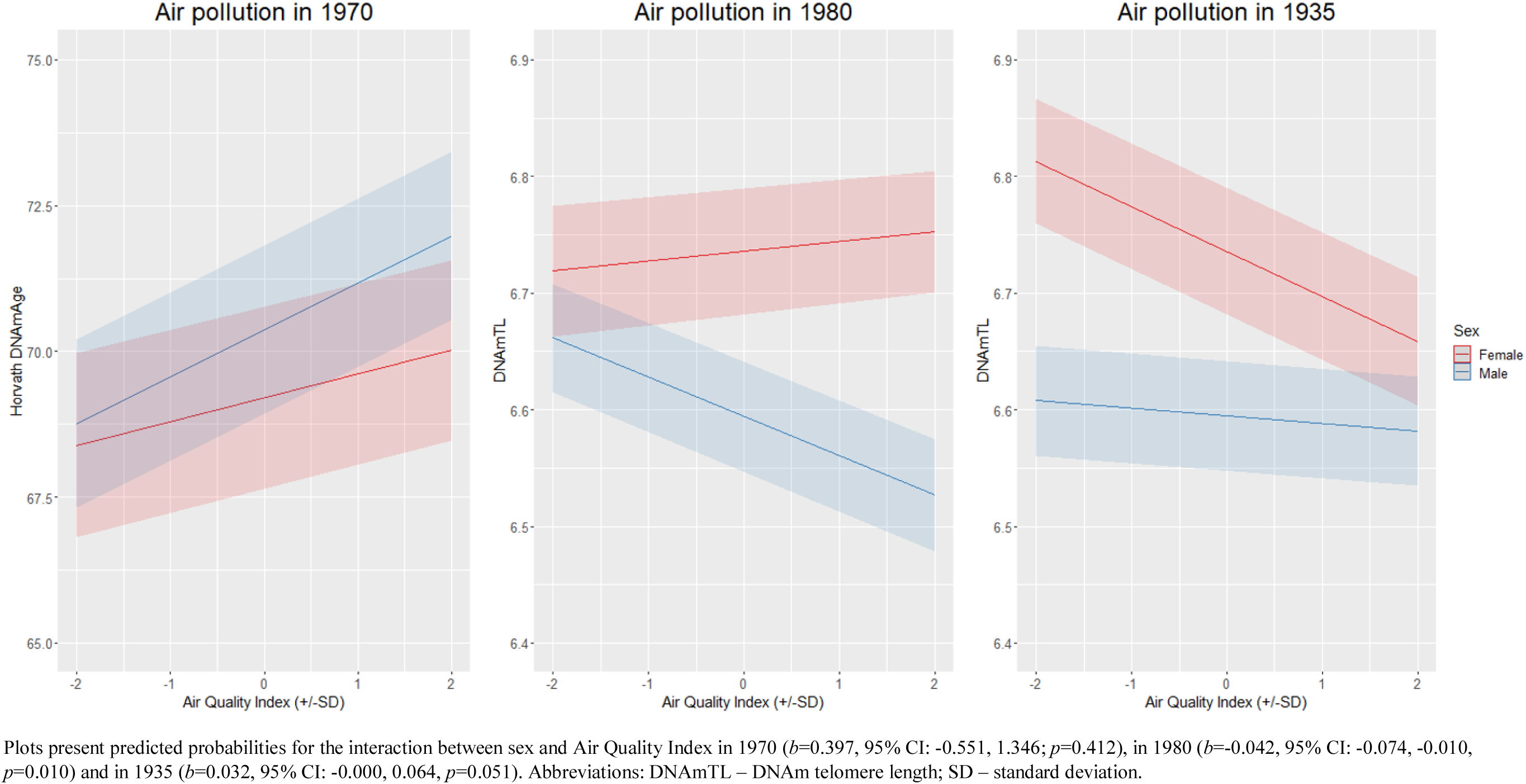
Exposure to air pollution in 1935, 1970 and 1980, and DNAm-based biomarkers among males and females

**Supplementary Table S5.** Exposure to air pollution and accelerated ageing in the Lothian Birth Cohort 1936 after adjusting for white blood cell proportions.

| Exposure | Total<br>(*n*=525, *obs*=1782)<br>Horvath DNAmAge<sup>a</sup><br>Air pollution in 1970 |  |  |  | Male<br>(*n*=277, *obs*=950)<br>DNAmTL<sup>b</sup><br>Air pollution in 1980 |  |  |  | Female<br>(*n*=248, *obs*=832)<br>DNAmTL<sup>c</sup><br>Air pollution in 1935 |  |  |  |
| --- | --- | --- | --- | --- | --- | --- | --- | --- | --- | --- | --- | --- |
| | Estimate | (95% CI) | *p* | $\beta$ | Estimate | (95% CI) | *p* | $\beta$ | Estimate | (95% CI) | *p* | $\beta$ |
| Air Quality Index (in 1 *SD*) | **0.580** | **0.148, 1.011** | **0.009** | **0.076** | **-0.029** | **-0.051, -0.007** | **0.009** | **-0.131** | **-0.032** | **-0.054, -0.011** | **0.004** | **-0.147** |
| Air pollutants (in 1 $\mu\text{g}/\text{m}^3$) | | | | | | | | | | | | |
| □PM<sub>2.5</sub> | 0.290 | 0.037, 0.544 | 0.025 | 0.065 | **-0.019** | **-0.034, -0.003** | **0.019** | **-0.118** | **-0.002** | **-0.004, -0.001** | **0.005** | **-0.144** |
| □SO<sub>2</sub> | 0.075 | 0.002, 0.148 | 0.046 | 0.059 | -0.004 | -0.008, 0.000 | 0.066 | -0.093 | **-0.001** | **-0.002, -0.000** | **0.004** | **-0.145** |
| □NO<sub>2</sub> | 0.109 | 0.005, 0.214 | 0.041 | 0.060 | **-0.007** | **-0.012, -0.002** | **0.007** | **-0.137** | **-0.005** | **-0.010, -0.001** | **0.011** | **-0.130** |
| □O<sub>3</sub> | -0.084 | -0.211, 0.043 | 0.195 | -0.038 | 0.006 | -0.000, 0.011 | 0.055 | 0.097 | **0.007** | **0.001, 0.013** | **0.017** | **0.122** |

Models were fitted with mixed-effects regression with random intercepts for study participants. Coefficients for the Air Quality Index are standardized, for air pollutants coefficients are expressed in $\mu\text{g}/\text{m}^3$ . We provide fully standardized coefficients to aid comparison. In addition to indicated confounders, all models are adjusted for white blood cell proportions using Houseman's algorithm (i.e. D8T, CD4T, NK, Bcell, Mono, Gran). Bold typeface denotes FDR-corrected significance. Abbreviations: DNAmTL – DNAm telomere length, SD – standard deviation.
<sup>a</sup>Models were adjusted for sex, age, parental occupational social class, childhood smoking and years spent in education;
<sup>b</sup>Models were adjusted for age, parental occupational social class, childhood smoking and years spent in education;
<sup>c</sup>Models were adjusted for age and parental occupational social class.

**Supplementary Table S6.** Exposure to air pollution and accelerated ageing in the Lothian Birth Cohort 1936 after controlling for batch effects.

| Exposure | Total<br>(*n*=525, *obs*=1782)<br>Horvath DNAmAge<sup>a</sup><br>Air pollution in 1970 |  |  |  | Male<br>(*n*=277, *obs*=950)<br>DNAmTL<sup>b</sup><br>Air pollution in 1980 |  |  |  | Female<br>(*n*=248, *obs*=832)<br>DNAmTL<sup>c</sup><br>Air pollution in 1935 |  |  |  |
| --- | --- | --- | --- | --- | --- | --- | --- | --- | --- | --- | --- | --- |
| | Estimate | 95% CI | *p* | $\beta$ | Estimate | 95% CI | *p* | $\beta$ | Estimate | 95% CI | *p* | $\beta$ |
| Air Quality Index (in 1 *SD*) | 0.602 | 0.132, 1.071 | 0.012 | 0.079 | **-0.036** | **-0.057, -0.014** | **0.001** | **-0.159** | **-0.035** | **-0.058, -0.012** | **0.003** | **-0.160** |
| Air pollutants (in 1 $\mu\text{g}/\text{m}^3$) | | | | | | | | | | | | |
| □PM<sub>2.5</sub> | 0.298 | 0.022, 0.574 | 0.035 | 0.067 | **-0.023** | **-0.038, -0.008** | **0.003** | **-0.146** | **-0.003** | **-0.004, -0.001** | **0.004** | **-0.155** |
| □SO<sub>2</sub> | 0.086 | 0.007, 0.166 | 0.034 | 0.068 | **-0.005** | **-0.009, -0.001** | **0.022** | **-0.114** | **-0.001** | **-0.002, -0.000** | **0.004** | **-0.155** |
| □NO<sub>2</sub> | 0.109 | -0.005, 0.224 | 0.062 | 0.059 | **-0.008** | **-0.012, -0.003** | **0.002** | **-0.151** | **-0.006** | **-0.010, -0.002** | **0.008** | **-0.143** |
| □O<sub>3</sub> | -0.095 | -0.233, 0.044 | 0.180 | -0.043 | **0.006** | **0.001, 0.012** | **0.033** | **0.106** | **0.008** | **0.002, 0.014** | **0.015** | **0.130** |

Models were fitted with mixed-effects regression with random intercepts for study participants and for batch variables (i.e. set of experiment, hybridisation date, array, position in array, plate).
Coefficients for the Air Quality Index are standardized, for air pollutants coefficients are expressed in $\mu\text{g}/\text{m}^3$ . Bold typeface denotes FDR-corrected significance. Abbreviations: DNAmTL – DNAm telomere length, SD – standard deviation.
<sup>a</sup>Models were adjusted for sex, age, parental occupational social class, childhood smoking and years spent in education;
<sup>b</sup>Models were adjusted for age, parental occupational social class, childhood smoking and years spent in education;
<sup>c</sup>Models were adjusted for age and parental occupational social class.

**Supplementary Table S7.**
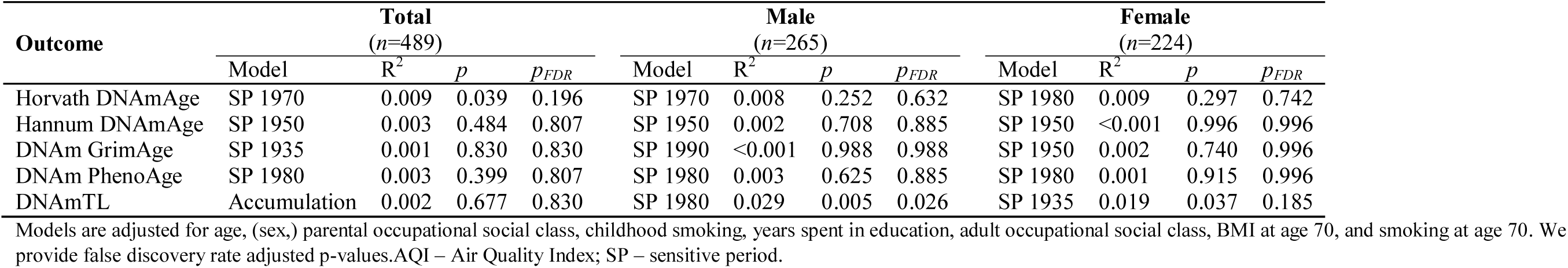
Selecting best-fit life-course models for the association between Air Quality Index and markers of biological ageing in the Lothian Birth Cohort 1936, after adjusting for all life course confounders.

| Outcome | Total<br>(n=489) |  |  |  | Male<br>(n=265) |  |  |  | Female<br>(n=224) |  |  |  |
| --- | --- | --- | --- | --- | --- | --- | --- | --- | --- | --- | --- | --- |
|  | Model | R<sup>2</sup> | p | p<sub>FDR</sub> | Model | R<sup>2</sup> | p | p<sub>FDR</sub> | Model | R<sup>2</sup> | p | p<sub>FDR</sub> |
| Horvath DNAmAge | SP 1970 | 0.009 | 0.039 | 0.196 | SP 1970 | 0.008 | 0.252 | 0.632 | SP 1980 | 0.009 | 0.297 | 0.742 |
| Hannum DNAmAge | SP 1950 | 0.003 | 0.484 | 0.807 | SP 1950 | 0.002 | 0.708 | 0.885 | SP 1950 | <0.001 | 0.996 | 0.996 |
| DNAm GrimAge | SP 1935 | 0.001 | 0.830 | 0.830 | SP 1990 | <0.001 | 0.988 | 0.988 | SP 1950 | 0.002 | 0.740 | 0.996 |
| DNAm PhenoAge | SP 1980 | 0.003 | 0.399 | 0.807 | SP 1980 | 0.003 | 0.625 | 0.885 | SP 1980 | 0.001 | 0.915 | 0.996 |
| DNAmTL | Accumulation | 0.002 | 0.677 | 0.830 | SP 1980 | 0.029 | 0.005 | 0.026 | SP 1935 | 0.019 | 0.037 | 0.185 |

Models are adjusted for age, (sex,) parental occupational social class, childhood smoking, years spent in education, adult occupational social class, BMI at age 70, and smoking at age 70. We provide false discovery rate adjusted p-values. AQI – Air Quality Index; SP – sensitive period.

**Supplementary Table S8.** Exposure to air pollution and accelerated ageing in the Lothian Birth Cohort 1936 after controlling for all confounders.

| Exposure | Total<br>(*n*=525, *obs*=1782)<br>Horvath DNAmAge<br>Air pollution in 1970 |  |  |  | Male<br>(*n*=277, *obs*=950)<br>DNAmTL<br>Air pollution in 1980 |  |  |  | Female<br>(*n*=248, *obs*=832)<br>DNAmTL<br>Air pollution in 1935 |  |  |  |
| --- | --- | --- | --- | --- | --- | --- | --- | --- | --- | --- | --- | --- |
|  | Estimate | 95% CI | p | β | Estimate | 95% CI | p | β | Estimate | 95% CI | p | β |
| Air Quality Index (in 1 *SD*) | 0.613 | 0.142, 1.084 | 0.011 | 0.081 | **-0.034** | **-0.055, -0.012** | **0.002** | **-0.151** | **-0.031** | **-0.054, -0.008** | **0.009** | **-0.141** |
| Air pollutants (in 1 $\mu\text{g}/\text{m}^3$) | | | | | | | | | | | | |
| □PM<sub>2.5</sub> | 0.292 | 0.016, 0.569 | 0.039 | 0.066 | **-0.022** | **-0.037, -0.007** | **0.004** | **-0.139** | **-0.002** | **-0.004, -0.001** | **0.011** | **-0.138** |
| □SO<sub>2</sub> | 0.087 | 0.007, 0.167 | 0.033 | 0.069 | **-0.005** | **-0.009, -0.000** | **0.031** | **-0.107** | **-0.001** | **-0.002, -0.000** | **0.010** | **-0.139** |
| □NO<sub>2</sub> | 0.109 | -0.006, 0.223 | 0.064 | 0.059 | **-0.007** | **-0.012, -0.002** | **0.003** | **-0.146** | **-0.005** | **-0.010, -0.001** | **0.023** | **-0.124** |
| □O<sub>3</sub> | -0.087 | -0.226, 0.052 | 0.222 | -0.040 | **0.006** | **0.000, 0.012** | **0.036** | **0.105** | **0.007** | **0.001, 0.013** | **0.034** | **0.115** |

Models were fitted with mixed-effects regression with random intercepts for study participants and adjusted for age, (sex,) parental occupational social class, childhood smoking, years spent in education, adult occupational social class, BMI and smoking at age 70. Coefficients for the Air Quality Index are standardized, for air pollutants coefficients are expressed in $\mu\text{g}/\text{m}^3$ . Bold typeface denotes FDR-corrected significance. Abbreviations: DNAmTL – DNAm telomere length, SD – standard deviation.

**Supplementary Table S9.**
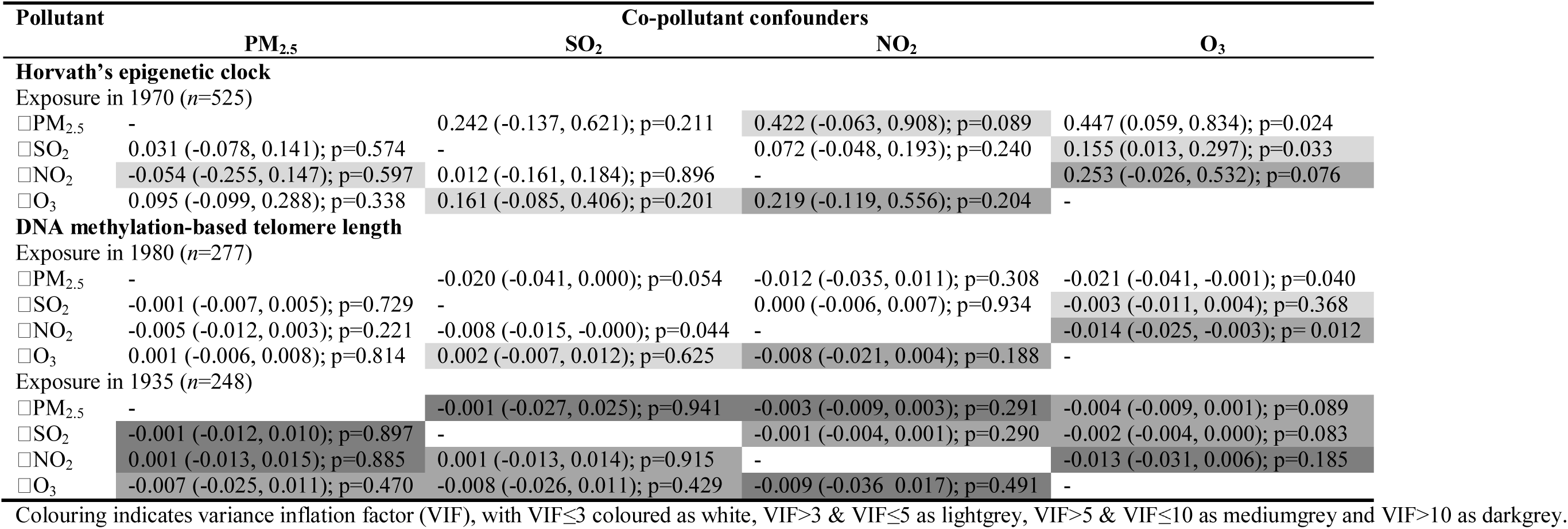
Two-pollutant models controlling for co-pollutant confounding

